# Statistical Analysis Plan for the Treatment of Cardiovascular disease with low dose Rivaroxaban in Advanced CKD (TRACK) trial

**DOI:** 10.1101/2025.11.25.25340065

**Authors:** Laurent Billot, Muh Geot Wong, An De Vriese, David Collister, Adrien Flahault, Lily Mushahar, Raja Ramachandran, Habib Skhiri, Ahmed Shaman, Jan Menne, Martin Gallagher

## Abstract

The Treatment of cardiovascular disease with low dose Rivaroxaban in Advanced Chronic Kidney Disease (TRACK) trial is a randomised quadruple-blind phase IV clinical trial to determine whether low-dose rivaroxaban (2.5 mg daily) reduces the risk of major adverse cardiovascular events compared to placebo. It aims to recruit approximately 2,000 patients with advanced chronic kidney disease (stages 4, 5 or dialysis-dependent) and an elevated cardiovascular risk.

This statistical analysis plan pre-specifies the method of analysis for every outcome and key variable collected in the trial. The primary outcome is the time from randomisation to first occurrence of major cardiovascular event including death from cardiovascular cause, myocardial infarction, stroke or peripheral artery disease event.

The primary analysis will consist in a Cox proportional hazard model adjusted for stratification variables. The analysis plan also includes planned sensitivity analyses including covariate adjustments and subgroup analyses.

## 3 Introduction

### 3.1 Study population

#### 3.1.1 Eligibility/Inclusion Criteria

[1] Age ≥18 years

[2] Kidney failure on haemodialysis or peritoneal dialysis, or chronic kidney disease stage 4 or 5 (eGFR ≤29 mL/min/1.73 m^2^) not receiving renal replacement therapy

[3] Elevated cardiovascular risk, defined by at least one of the following:

- Coronary artery disease
- Non-haemorrhagic non-lacunar stroke
- Peripheral artery disease
- Diabetes mellitus
- Age ≥65 years

#### 3.1.2 Exclusion criteria

[1] Mechanical/prosthetic heart valve (does not include bioprosthetic valves that do not require therapeutic anticoagulation)

[2] Indication for, or contraindication to, anticoagulant therapy

[3] High bleeding risk including any coagulopathy

[4] Lesion or condition considered to be a significant risk of major bleeding

[5] Major bleeding episode in the 30 days prior to study enrolment, or any active and clinically significant bleeding

[6] Current treatment with P2Y_12_ inhibitors/adenosine diphosphate receptor inhibitors (clopidogrel, prasugrel, ticagrelor, cangrelor) or phosphodiesterase inhibitors (dipyridamole), where the treating physician or patient does not wish to stop these medications

[7] Concurrent treatment with strong inhibitors of combined CYP3A4 and P-glycoprotein; or strong inducers of CYP3A4

[8] Any stroke within 1 month prior to enrolment

[9] Any previous history of a hemorrhagic or lacunar stroke

[10] Severe heart failure with known ejection fraction <30% or NYHA class III or IV symptoms

[11] History of hypersensitivity or known contraindication to rivaroxaban

[12] Uncontrolled hypertension (systolic BP ≥180 mm Hg or diastolic BP ≥110 mm Hg) at the time of screening

[13] Haemoglobin <90 g/L, or platelet count <100 x10^9^/L

[14] Significant liver disease (defined as Child-Pugh Class B or C) or ALT >3 times upper normal limit,

[15] Kidney transplant recipients with a functioning allograft, or scheduled for living-donor kidney transplant surgery,

[16] All countries except Europe: Pregnancy or intention to become pregnant or breast-feeding, Europe only: Women who are not in a postmenopausal state, where postmenopausal is defined as no menses for 12 months without alternative medical causes,

[17] Inability to understand or comply with the requirements of the study

### 3.2 Interventions

Following a placebo run-in phase, eligible patients with at least 80% adherence were randomly allocated to receive either rivaroxaban 2.5 mg twice daily or matched placebo twice daily.

### 3.3 Outcomes

#### 3.3.1 Primary efficacy outcome

The primary efficacy outcome is a is a major adverse cardiovascular events (MACE) composite, defined as the time from randomisation to first occurrence of death from cardiovascular causes, myocardial infarction, stroke or peripheral artery disease event.

#### 3.3.2 Secondary efficacy outcomes

Secondary endpoints include the following:

1. Composite of cardiovascular death, myocardial infarction or stroke
2. All-cause death
3. Composite of all-cause death, myocardial infarction or stroke
4. Composite of all-cause death, myocardial infarction, stroke or peripheral artery disease event
5. Cardiovascular death
6. Stroke
7. Myocardial infarction
8. Peripheral artery disease event
9. Venous thromboembolism

#### 3.3.3 Tertiary efficacy outcomes

1. Thrombosis of dialysis vascular access among participants with an AV fistula or graft (including those receiving haemodialysis, peritoneal dialysis or people with CKD stage 4/5)
2. Self-reported EQ-5D-5L
3. Health care resource utilisation and costs

#### 3.3.4 Safety outcomes

1. Modified ISTH major bleeding, defined as:

- Fatal bleeding, and/or
- Symptomatic bleeding in a critical area or organ, such as intracranial, intraspinal, intraocular, retroperitoneal, intraarticular or pericardial, or intramuscular with compartment syndrome, or bleeding into the surgical site requiring reoperation, and/or
- Bleeding leading to hospitalisation.
2. Gastrointestinal bleeding
3. Serious adverse events

#### 3.3.5 Net clinical benefit outcome

Net clinical benefit includes major efficacy and safety outcomes and is defined as a composite of cardiovascular death, myocardial infarction, stroke, peripheral artery disease events, fatal bleeding or symptomatic bleeding into a critical organ.

### 3.4 Randomisation and blinding

Eligible patients are randomised via a web-based system via a password-protected encrypted website to either low dose rivaroxaban (2.5 mg twice daily), or placebo, in a 1:1 ratio. A covariate-balancing adaptive allocation algorithm is used to minimise imbalance across treatment groups in the following variables: (1) study site, (2) use of aspirin at the time of randomization (yes or no), (3) CKD stage (CKD stage 4/5 not receiving renal replacement therapy or dialysis-dependent kidney failure), and (4) presence of diabetes mellitus. Trial participants, site staff, outcome assessors and investigators are blinded to an individual’s randomised allocation. Analyses outlined in this analysis plan will be programmed and validated using randomly-shuffled treatment allocations. Following database lock and the validation of analyses, programs will be rerun using the real treatment allocations.

### 3.5 Sample size

Assuming a primary endpoint event rate of 10 per 100 person-years in the control group and a two-sided alpha of 4.82% (adjusted for interim analyses – see Section 4.1.1), a total of 515 participants experiencing a primary outcome provides 90% power to detect a 25% risk reduction (hazard ratio of 0.75), or 80% power to detect a 22% risk reduction (hazard ratio 0.78) with the study intervention (low dose rivaroxaban) versus placebo using an intention to treat approach.

The study planned to continue until a total of 515 participants experienced a primary endpoint event, recruiting approximately 1900 participants over 3 years with a maximum duration of follow-up of 5 years from the recruitment of the first participant (i.e. a median follow-up of 3.5 years assuming uniform recruitment rates). Following a recommendation from the DSMB, the trial was stopped early with 1,463 participants randomised.

## 4 Statistical analyses

### 4.1 General principles

#### 4.1.1 Level of statistical significance

One formal efficacy interim analysis was conducted in October 2024 and included 181 patients with a primary outcome event. The study protocol and DSMB charter pre-specified the use of Haybittle-Peto [1] boundaries to guide early stopping for efficacy while maintaining the overall type-I error to 5% (two-sided). With one interim analysis performed at 181 events (0.27% of alpha spent), the final significance level is therefore set at 4.86% (two-sided) corresponding to a z-value of 1.972.

Given the clear outcome hierarchy, the analysis of the primary outcome will not be adjusted for multiplicity (aside from the one arising from interim analyses). Sensitivity analyses of the primary outcome will also not adjust for multiplicity as they are conducted to assess the robustness of the main findings using different approaches/assumptions.

For the 9 secondary outcomes, we will use the following approach:

1. For all-cause death and the four components of the primary outcome (cardiovascular death, stroke, myocardial infarction and peripheral artery disease events), the family-wise error rate will be controlled using a hierarchical step-down approach with a Holm-Sidak correction [2]. Briefly, the Holm-Sidak approach consists of ordering all p-values from smallest to largest, and then comparing them to an adjusted level of significance calculated as 1-(1-0.05)^1/C^, where C indicates the number of comparisons that remain. With 5 endpoints, the smallest p value will be compared to 1-(1-0.05)^1/5^, the second p value to 1-(1-0.05)^1/4^, with the last one being compared to 1-(1-0.05) (i.e. 0.05). The sequential testing procedure stops as soon as a p value fails to reach the corrected significance level.
2. For the 5 remaining secondary outcomes including the alternative composite outcomes, no formal test will be conducted and we will only report point estimates and 95% confidence intervals to assess consistency of treatment effects.

For other outcomes including safety outcomes and other analyses (e.g. win ratio) we will not adjust the significance level. The results will be considered as exploratory. For the net clinical benefit outcome, we will keep the significance level at 5% given it includes key safety outcomes and we wish to be able to detect a potential signal.

#### 4.1.2 Analysis populations

All analyses will be performed in the intention to treat (ITT) analysis set. Sensitivity analyses for selected outcomes (as flagged in the relevant sections of the SAP) will be conducted in the per-protocol analysis set.

The ***intention-to-treat (ITT) analysis set*** will include all randomised patients who were eligible. Patients will be analysed according to the group they were randomised to, regardless of compliance or post-randomisation events. The ITT set will be used to assess all efficacy and safety outcomes.

The ***per-protocol (PP) analysis set*** will be defined as patients who received the randomised treatment at least once and did not have any discontinuation of the study treatment. The PP set will be used for sensitivity analyses of the primary outcome.

The main efficacy (primary and secondary outcomes) and safety analyses (bleeding outcomes) will be repeated in the following subsets of patients regardless of results:

1. Patients receiving aspirin at baseline
2. Patients not receiving aspirin at baseline
3. Patients on dialysis at baseline
4. Patients not on dialysis at baseline

#### 4.1.3 Missing data handling

Except where specifically indicated, analyses will be conducted using all available data with no imputation. Given the nature of the primary endpoint (time to major cardiovascular outcome), there is no reasonable way to assess whether some events are potentially missing (ie. unreported). We will therefore perform the analysis assuming that all events that occurred were reported, and without imputation of missing data. All reported events will be included in the time-to-first-event analysis with every participant censored at the known time of event, or the time when the participant was last known to be alive and free of an event. The same approach will be used for every survival endpoint.

#### 4.1.4 Adjustment for stratification variables

Randomisation is stratified by four variables: (1) study site, (2) use of aspirin at the time of randomisation, (3) CKD stage (non-dialysis CKD stage 4/5 vs. dialysis-dependent kidney failure), and (4) presence or absence of diabetes mellitus. By default, all analyses will be adjusted by these four variables with site as a random effect and the three others as fixed effects. Further covariate adjustments will be specifically indicated where relevant.

#### 4.1.5 Statistical software

Analyses will be conducted primarily using SAS (version 9.3 or above) or R (version R-4.3.1 or above).

### 4.2 Subject disposition

Overall subject disposition and flow will be reported using a Consort diagram [3]. In addition, we will summarise details by visit and randomised arm including visit types (clinic, telephone, etc.) and patient status (still in study, lost to follow-up, dead, etc.).

### 4.3 Analysis of baseline data

Socio-demographic characteristics, medical history and concomitant medications at the time of randomisation (baseline) will be summarised by treatment arm and overall using standard summary statistics i.e. n, mean, standard-deviation, median, quartiles, minimum and maximum for continuous variables and numerator, denominator and percentage for categorical variables. No statistical test will be performed; however, we will calculate standardised differences to quantify the degree of imbalance between randomised arms.

### 4.4 Analysis of compliance and dialysis details

Medication compliance will be reported using the following metrics:

- Tablet count
- Proportion of patients still on randomised therapy by visit
- Number and percentage of discontinuations by type (permanent or temporary) and reason
- Protocol deviations

Change in dialysis modalities over the course of the study will be summarised as follows:

- Number and proportion on dialysis (any modality) at each visit
- Number and proportion of patients who started or stopped dialysis after randomisation
- Number and proportion of patients who switched from haemodialysis to peritoneal dialysis at any point
- Number and proportion of patients who switched from peritoneal to haemodialysis dialysis at any point
- Number and proportion of patients who underwent kidney transplantation

These will be analysed descriptively only with no formal statistical test.

**Table 1.** Participant disposition.

|  | **Rivaroxaban**<br>(N = XXX) | **Placebo**<br>(N = XXX) | **Total**<br>(N = XXX) |
| --- | --- | --- | --- |
| **Randomised** | XX | XX | XX |
| Australia | XX (XX%) | XX (XX%) | XX (XX%) |
| Belgium | XX (XX%) | XX (XX%) | XX (XX%) |
| Canada | XX (XX%) | XX (XX%) | XX (XX%) |
| France | XX (XX%) | XX (XX%) | XX (XX%) |
| Germany | XX (XX%) | XX (XX%) | XX (XX%) |
| India | XX (XX%) | XX (XX%) | XX (XX%) |
| Malaysia | XX (XX%) | XX (XX%) | XX (XX%) |
| Nepal | XX (XX%) | XX (XX%) | XX (XX%) |
| Saudi Arabia | XX (XX%) | XX (XX%) | XX (XX%) |
| Singapore | XX (XX%) | XX (XX%) | XX (XX%) |
| Taiwan | XX (XX%) | XX (XX%) | XX (XX%) |
| Tunisia | XX (XX%) | XX (XX%) | XX (XX%) |
| **Clinic visit attendance** |  |  |  |
| Month 1 | XX (XX%) | XX (XX%) | XX (XX%) |
| Month 3 | XX (XX%) | XX (XX%) | XX (XX%) |
| Month 6 | XX (XX%) | XX (XX%) | XX (XX%) |
| Month 9 | XX (XX%) | XX (XX%) | XX (XX%) |
| Etc. |  |  |  |
| Month 60 | XX (XX%) | XX (XX%) | XX (XX%) |
| End of study | XX (XX%) | XX (XX%) | XX (XX%) |
| **Trial withdrawal** |  |  |  |
| Death | XX (XX%) | XX (XX%) | XX (XX%) |
| Consent withdrawn | XX (XX%) | XX (XX%) | XX (XX%) |
| Hospital transfer | XX (XX%) | XX (XX%) | XX (XX%) |
| Lost to follow-up | XX (XX%) | XX (XX%) | XX (XX%) |
| Other | XX (XX%) | XX (XX%) | XX (XX%) |

**Table 2.**
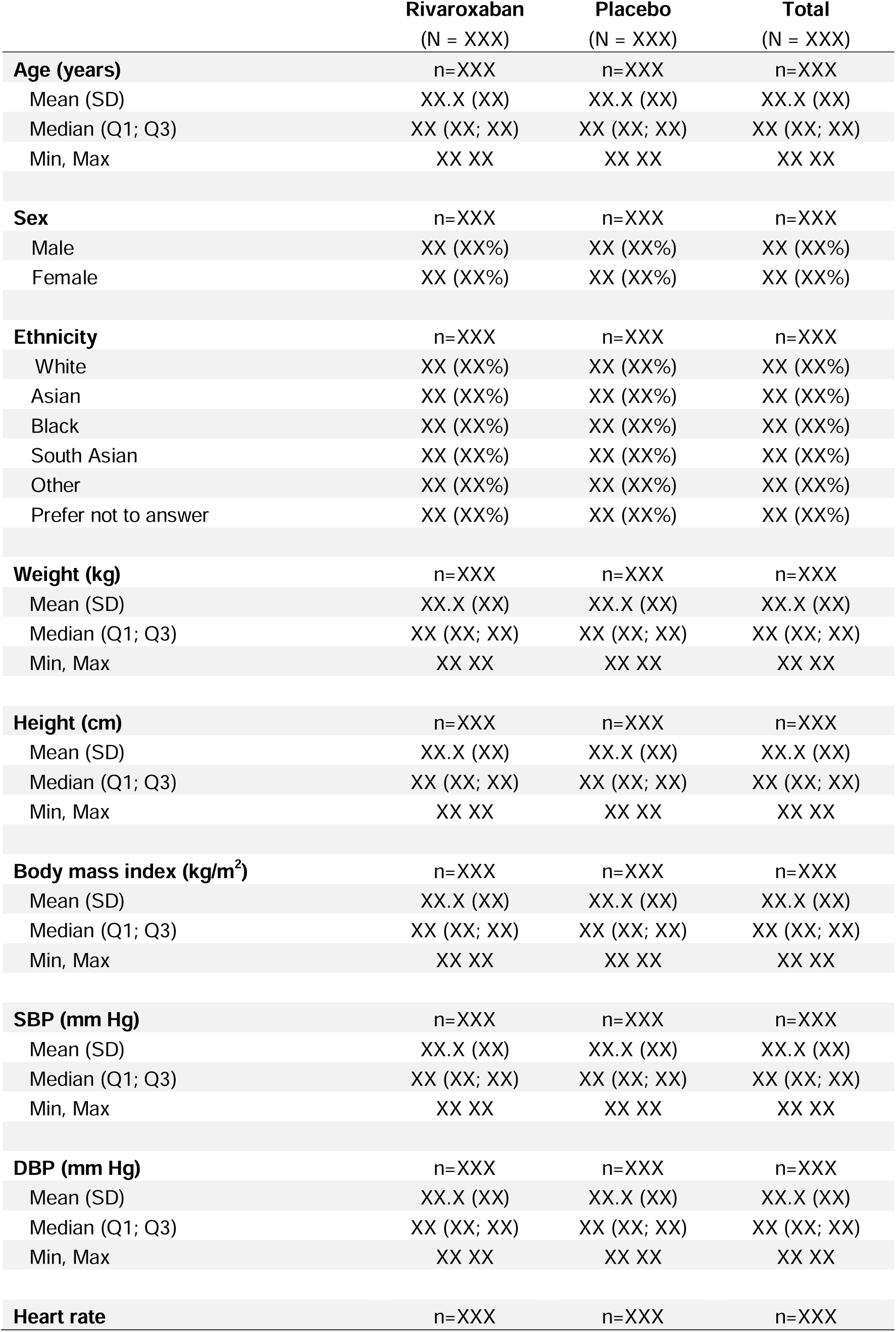
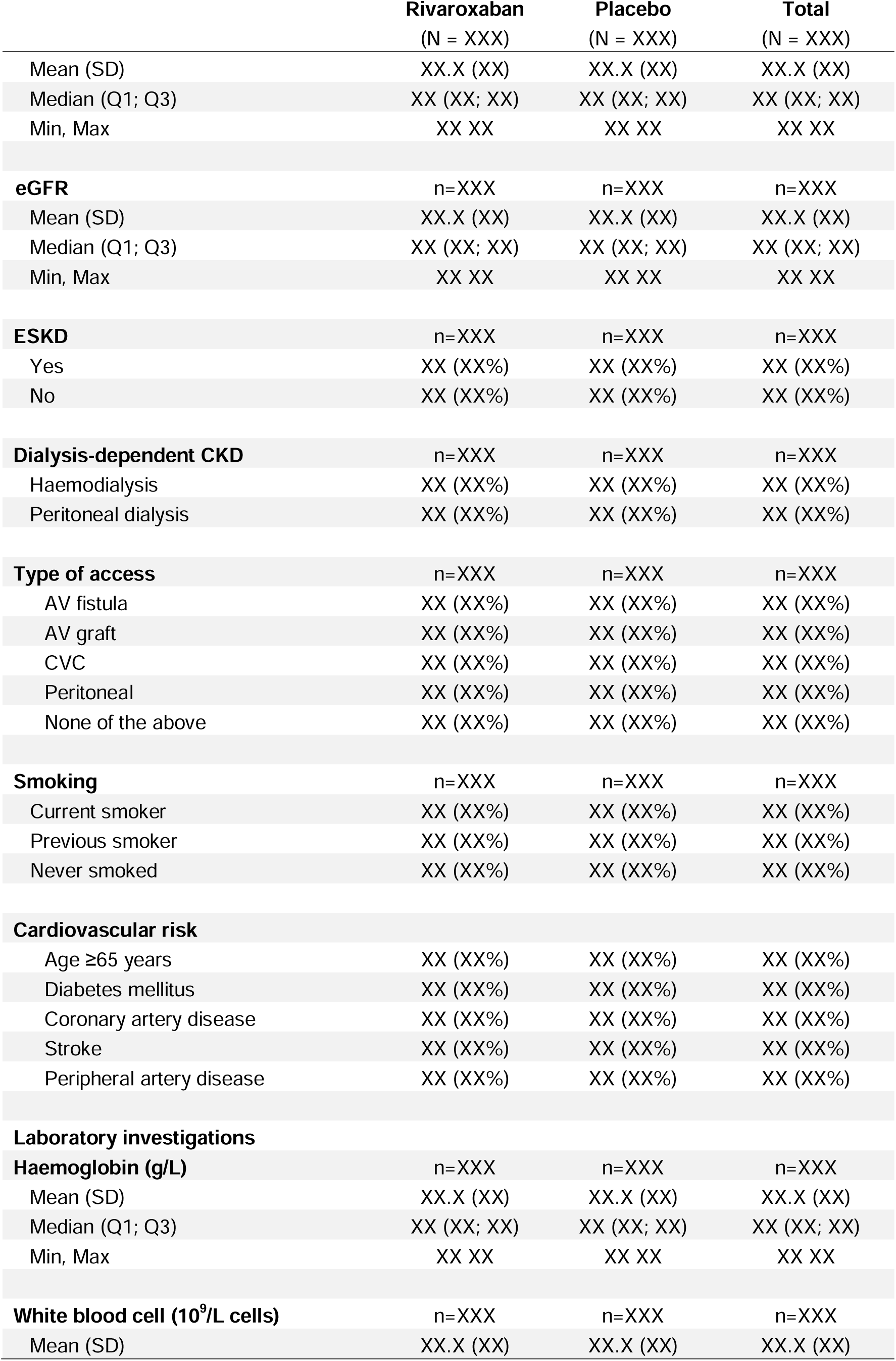
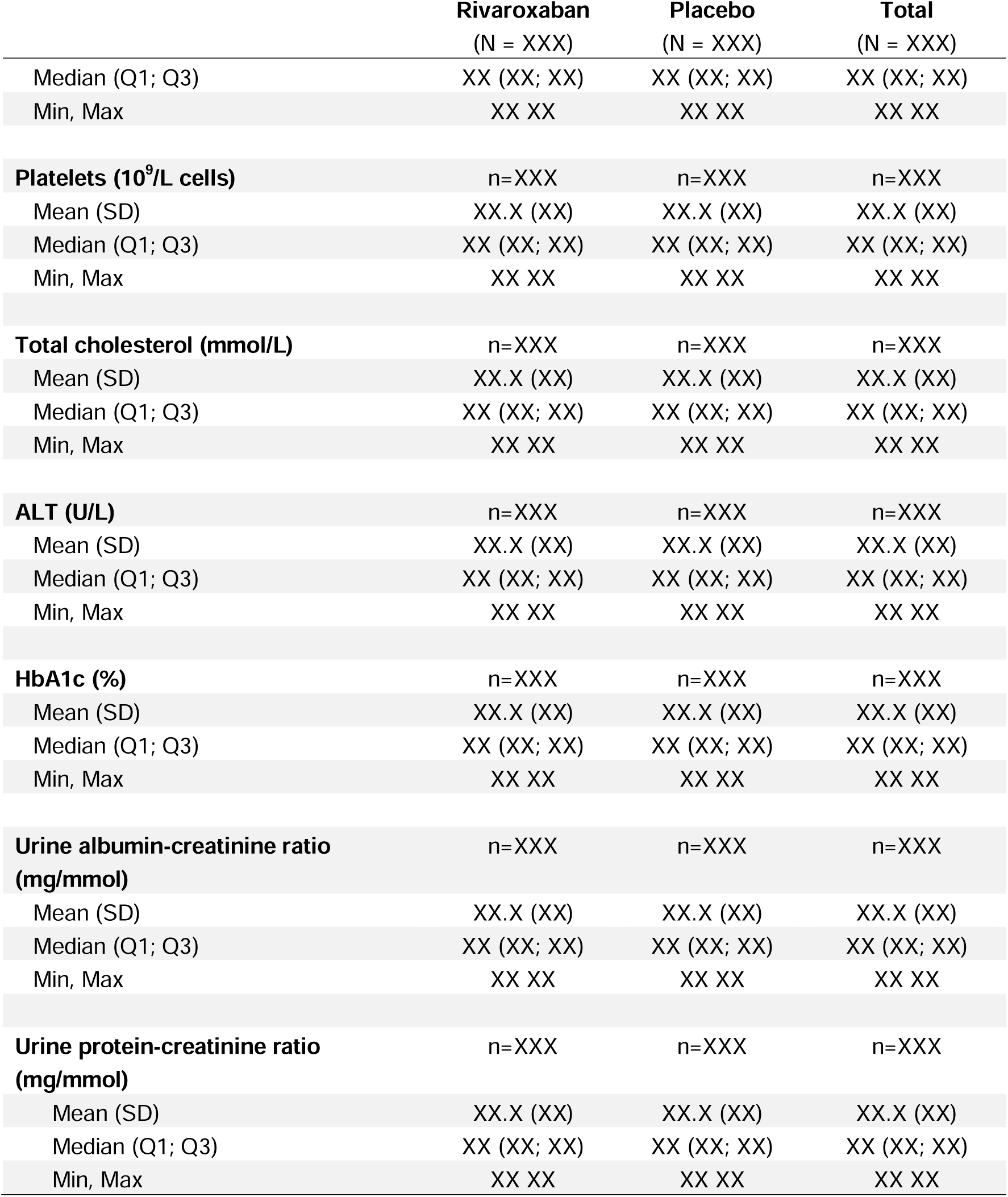
Key baseline characteristics.

|  | **Rivaroxaban**<br>(N = XXX) | **Placebo**<br>(N = XXX) | **Total**<br>(N = XXX) |
| --- | --- | --- | --- |
| **Age (years)** | n=XXX | n=XXX | n=XXX |
| Mean (SD) | XX.X (XX) | XX.X (XX) | XX.X (XX) |
| Median (Q1; Q3) | XX (XX; XX) | XX (XX; XX) | XX (XX; XX) |
| Min, Max | XX XX | XX XX | XX XX |
| **Sex** | n=XXX | n=XXX | n=XXX |
| Male | XX (XX%) | XX (XX%) | XX (XX%) |
| Female | XX (XX%) | XX (XX%) | XX (XX%) |
| **Ethnicity** | n=XXX | n=XXX | n=XXX |
| White | XX (XX%) | XX (XX%) | XX (XX%) |
| Asian | XX (XX%) | XX (XX%) | XX (XX%) |
| Black | XX (XX%) | XX (XX%) | XX (XX%) |
| South Asian | XX (XX%) | XX (XX%) | XX (XX%) |
| Other | XX (XX%) | XX (XX%) | XX (XX%) |
| Prefer not to answer | XX (XX%) | XX (XX%) | XX (XX%) |
| **Weight (kg)** | n=XXX | n=XXX | n=XXX |
| Mean (SD) | XX.X (XX) | XX.X (XX) | XX.X (XX) |
| Median (Q1; Q3) | XX (XX; XX) | XX (XX; XX) | XX (XX; XX) |
| Min, Max | XX XX | XX XX | XX XX |
| **Height (cm)** | n=XXX | n=XXX | n=XXX |
| Mean (SD) | XX.X (XX) | XX.X (XX) | XX.X (XX) |
| Median (Q1; Q3) | XX (XX; XX) | XX (XX; XX) | XX (XX; XX) |
| Min, Max | XX XX | XX XX | XX XX |
| **Body mass index (kg/m<sup>2</sup>)** | n=XXX | n=XXX | n=XXX |
| Mean (SD) | XX.X (XX) | XX.X (XX) | XX.X (XX) |
| Median (Q1; Q3) | XX (XX; XX) | XX (XX; XX) | XX (XX; XX) |
| Min, Max | XX XX | XX XX | XX XX |
| **SBP (mm Hg)** | n=XXX | n=XXX | n=XXX |
| Mean (SD) | XX.X (XX) | XX.X (XX) | XX.X (XX) |
| Median (Q1; Q3) | XX (XX; XX) | XX (XX; XX) | XX (XX; XX) |
| Min, Max | XX XX | XX XX | XX XX |
| **DBP (mm Hg)** | n=XXX | n=XXX | n=XXX |
| Mean (SD) | XX.X (XX) | XX.X (XX) | XX.X (XX) |
| Median (Q1; Q3) | XX (XX; XX) | XX (XX; XX) | XX (XX; XX) |
| Min, Max | XX XX | XX XX | XX XX |
| **Heart rate** | n=XXX | n=XXX | n=XXX |

|  | **Rivaroxaban** | **Placebo** | **Total** |
| --- | --- | --- | --- |
|  | (N = XXX) | (N = XXX) | (N = XXX) |
| Mean (SD) | XX.X (XX) | XX.X (XX) | XX.X (XX) |
| Median (Q1; Q3) | XX (XX; XX) | XX (XX; XX) | XX (XX; XX) |
| Min, Max | XX XX | XX XX | XX XX |
| **eGFR** | n=XXX | n=XXX | n=XXX |
| Mean (SD) | XX.X (XX) | XX.X (XX) | XX.X (XX) |
| Median (Q1; Q3) | XX (XX; XX) | XX (XX; XX) | XX (XX; XX) |
| Min, Max | XX XX | XX XX | XX XX |
| **ESKD** | n=XXX | n=XXX | n=XXX |
| Yes | XX (XX%) | XX (XX%) | XX (XX%) |
| No | XX (XX%) | XX (XX%) | XX (XX%) |
| **Dialysis-dependent CKD** | n=XXX | n=XXX | n=XXX |
| Haemodialysis | XX (XX%) | XX (XX%) | XX (XX%) |
| Peritoneal dialysis | XX (XX%) | XX (XX%) | XX (XX%) |
| **Type of access** | n=XXX | n=XXX | n=XXX |
| AV fistula | XX (XX%) | XX (XX%) | XX (XX%) |
| AV graft | XX (XX%) | XX (XX%) | XX (XX%) |
| CVC | XX (XX%) | XX (XX%) | XX (XX%) |
| Peritoneal | XX (XX%) | XX (XX%) | XX (XX%) |
| None of the above | XX (XX%) | XX (XX%) | XX (XX%) |
| **Smoking** | n=XXX | n=XXX | n=XXX |
| Current smoker | XX (XX%) | XX (XX%) | XX (XX%) |
| Previous smoker | XX (XX%) | XX (XX%) | XX (XX%) |
| Never smoked | XX (XX%) | XX (XX%) | XX (XX%) |
| **Cardiovascular risk** |  |  |  |
| Age ≥65 years | XX (XX%) | XX (XX%) | XX (XX%) |
| Diabetes mellitus | XX (XX%) | XX (XX%) | XX (XX%) |
| Coronary artery disease | XX (XX%) | XX (XX%) | XX (XX%) |
| Stroke | XX (XX%) | XX (XX%) | XX (XX%) |
| Peripheral artery disease | XX (XX%) | XX (XX%) | XX (XX%) |
| **Laboratory investigations** |  |  |  |
| **Haemoglobin (g/L)** | n=XXX | n=XXX | n=XXX |
| Mean (SD) | XX.X (XX) | XX.X (XX) | XX.X (XX) |
| Median (Q1; Q3) | XX (XX; XX) | XX (XX; XX) | XX (XX; XX) |
| Min, Max | XX XX | XX XX | XX XX |
| **White blood cell (10<sup>9</sup>/L cells)** | n=XXX | n=XXX | n=XXX |
| Mean (SD) | XX.X (XX) | XX.X (XX) | XX.X (XX) |
| Median (Q1; Q3) | XX (XX; XX) | XX (XX; XX) | XX (XX; XX) |
| Min, Max | XX XX | XX XX | XX XX |
| **Platelets (10<sup>9</sup>/L cells)** | n=XXX | n=XXX | n=XXX |
| Mean (SD) | XX.X (XX) | XX.X (XX) | XX.X (XX) |
| Median (Q1; Q3) | XX (XX; XX) | XX (XX; XX) | XX (XX; XX) |
| Min, Max | XX XX | XX XX | XX XX |
| **Total cholesterol (mmol/L)** | n=XXX | n=XXX | n=XXX |
| Mean (SD) | XX.X (XX) | XX.X (XX) | XX.X (XX) |
| Median (Q1; Q3) | XX (XX; XX) | XX (XX; XX) | XX (XX; XX) |
| Min, Max | XX XX | XX XX | XX XX |
| **ALT (U/L)** | n=XXX | n=XXX | n=XXX |
| Mean (SD) | XX.X (XX) | XX.X (XX) | XX.X (XX) |
| Median (Q1; Q3) | XX (XX; XX) | XX (XX; XX) | XX (XX; XX) |
| Min, Max | XX XX | XX XX | XX XX |
| **HbA1c (%)** | n=XXX | n=XXX | n=XXX |
| Mean (SD) | XX.X (XX) | XX.X (XX) | XX.X (XX) |
| Median (Q1; Q3) | XX (XX; XX) | XX (XX; XX) | XX (XX; XX) |
| Min, Max | XX XX | XX XX | XX XX |
| **Urine albumin-creatinine ratio**<br>**(mg/mmol)** | n=XXX | n=XXX | n=XXX |
| Mean (SD) | XX.X (XX) | XX.X (XX) | XX.X (XX) |
| Median (Q1; Q3) | XX (XX; XX) | XX (XX; XX) | XX (XX; XX) |
| Min, Max | XX XX | XX XX | XX XX |
| **Urine protein-creatinine ratio**<br>**(mg/mmol)** | n=XXX | n=XXX | n=XXX |
| Mean (SD) | XX.X (XX) | XX.X (XX) | XX.X (XX) |
| Median (Q1; Q3) | XX (XX; XX) | XX (XX; XX) | XX (XX; XX) |
| Min, Max | XX XX | XX XX | XX XX |

**Table 3.**
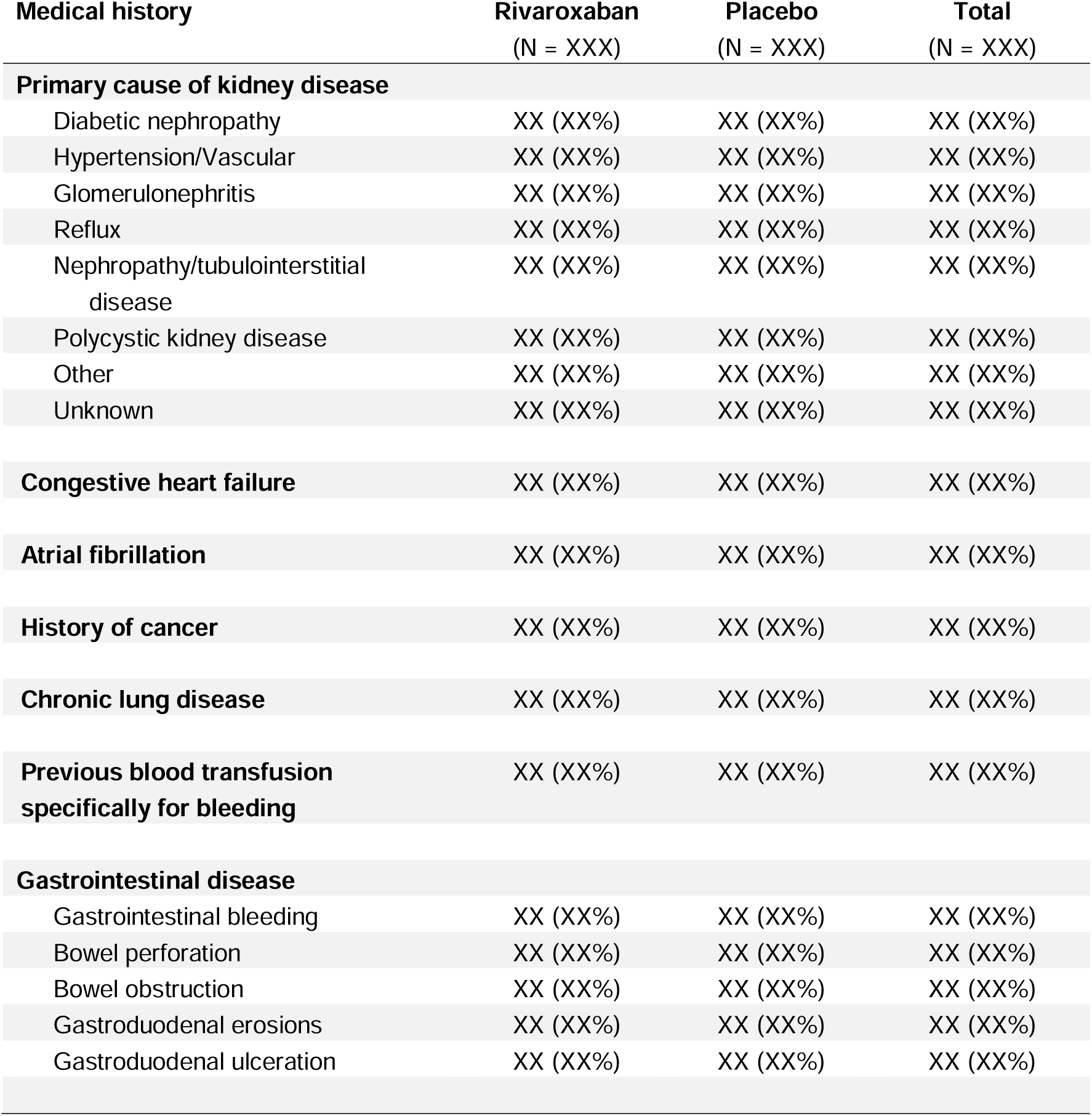
Medical history at baseline.

**Table 4.**
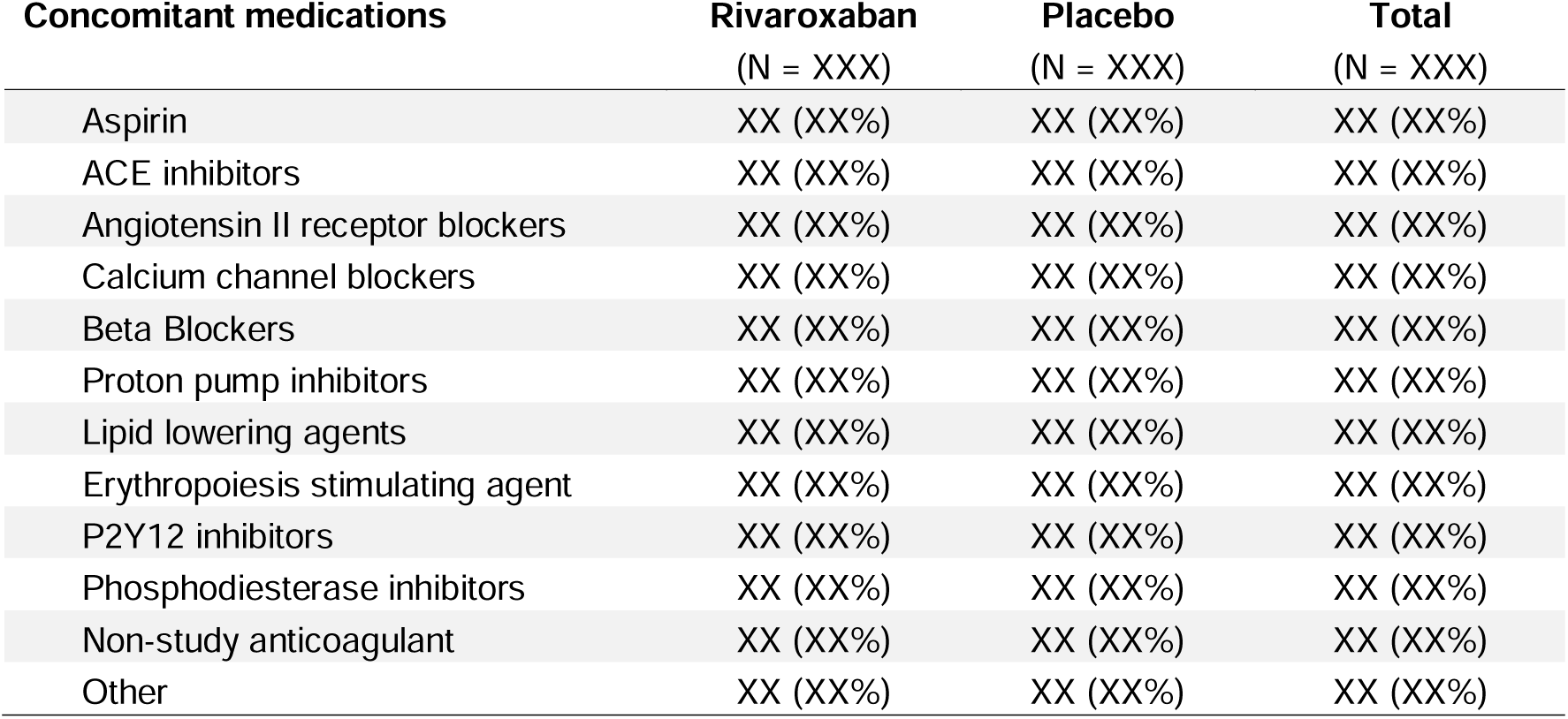
Concomitant medications at baseline.

**Table 5.** Study drug discontinuations.

| Type of discontinuation | Rivaroxaban<br>(N=XXXX) |  | Placebo<br>(N=XXXX) |  | Total<br>(N=XXXX) |  |
| --- | --- | --- | --- | --- | --- | --- |
|  | Event<br>s | Subjects | Event<br>s | Subjects | Event<br>s | Subjects |
| **Short-term discontinuation** | XXX | XX (XX%) | XXX | XX (XX%) | XXX | XX (XX%) |
| Invasive procedure | XXX | XX (XX%) | XXX | XX (XX%) | XXX | XX (XX%) |
| Short-term anticoagulation | XXX | XX (XX%) | XXX | XX (XX%) | XXX | XX (XX%) |
| Contraindicated medication | XXX | XX (XX%) | XXX | XX (XX%) | XXX | XX (XX%) |
| Condition or illness requiring<br>cessation | XXX | XX (XX%) | XXX | XX (XX%) | XXX | XX (XX%) |
| Other | XXX | XX (XX%) | XXX | XX (XX%) | XXX | XX (XX%) |
| **Long-term discontinuation** | XXX | XX (XX%) | XXX | XX (XX%) | XXX | XX (XX%) |
| Life-threatening or<br>intracranial bleeding | XXX | XX (XX%) | XXX | XX (XX%) | XXX | XX (XX%) |
| Long-term anticoagulation | XXX | XX (XX%) | XXX | XX (XX%) | XXX | XX (XX%) |
| Long-term DAPT | XXX | XX (XX%) | XXX | XX (XX%) | XXX | XX (XX%) |
| Contraindicated medication | XXX | XX (XX%) | XXX | XX (XX%) | XXX | XX (XX%) |
| Kidney transplantation | XXX | XX (XX%) | XXX | XX (XX%) | XXX | XX (XX%) |
| Pregnancy | XXX | XX (XX%) | XXX | XX (XX%) | XXX | XX (XX%) |
| Clinical decision by<br>physician | XXX | XX (XX%) | XXX | XX (XX%) | XXX | XX (XX%) |
| Participant request | XXX | XX (XX%) | XXX | XX (XX%) | XXX | XX (XX%) |
| Other | XXX | XX (XX%) | XXX | XX (XX%) | XXX | XX (XX%) |

**Table 6.** Adherence to study medication.

|  | **Rivaroxaban**<br>(N = XXX) | **Placebo**<br>(N = XXX) | **Total**<br>(N = XXX) |
| --- | --- | --- | --- |
| **Any missed tablets since last visit?** |  |  |  |
| **Visit XX** | XX/XXX (XX%) | XX/XXX (XX%) | XX/XXX (XX%) |
| **Visit XX** | XX/XXX (XX%) | XX/XXX (XX%) | XX/XXX (XX%) |
| **Etc. for all visits** |  |  |  |
| **Number of tablets remaining** |  |  |  |
| **Visit XX** | n=XXX | n=XXX | n=XXX |
| Mean (SD) | XX.X (XX) | XX.X (XX) | XX.X (XX) |
| Median (Q1; Q3) | XX (XX; XX) | XX (XX; XX) | XX (XX; XX) |
| Min, Max | XX XX | XX XX | XX XX |
| **Visit XX** | n=XXX | n=XXX | n=XXX |
| Mean (SD) | XX.X (XX) | XX.X (XX) | XX.X (XX) |
| Median (Q1; Q3) | XX (XX; XX) | XX (XX; XX) | XX (XX; XX) |
| Min, Max | XX XX | XX XX | XX XX |
| **Etc. for all visits** |  |  |  |

**Table 7.**
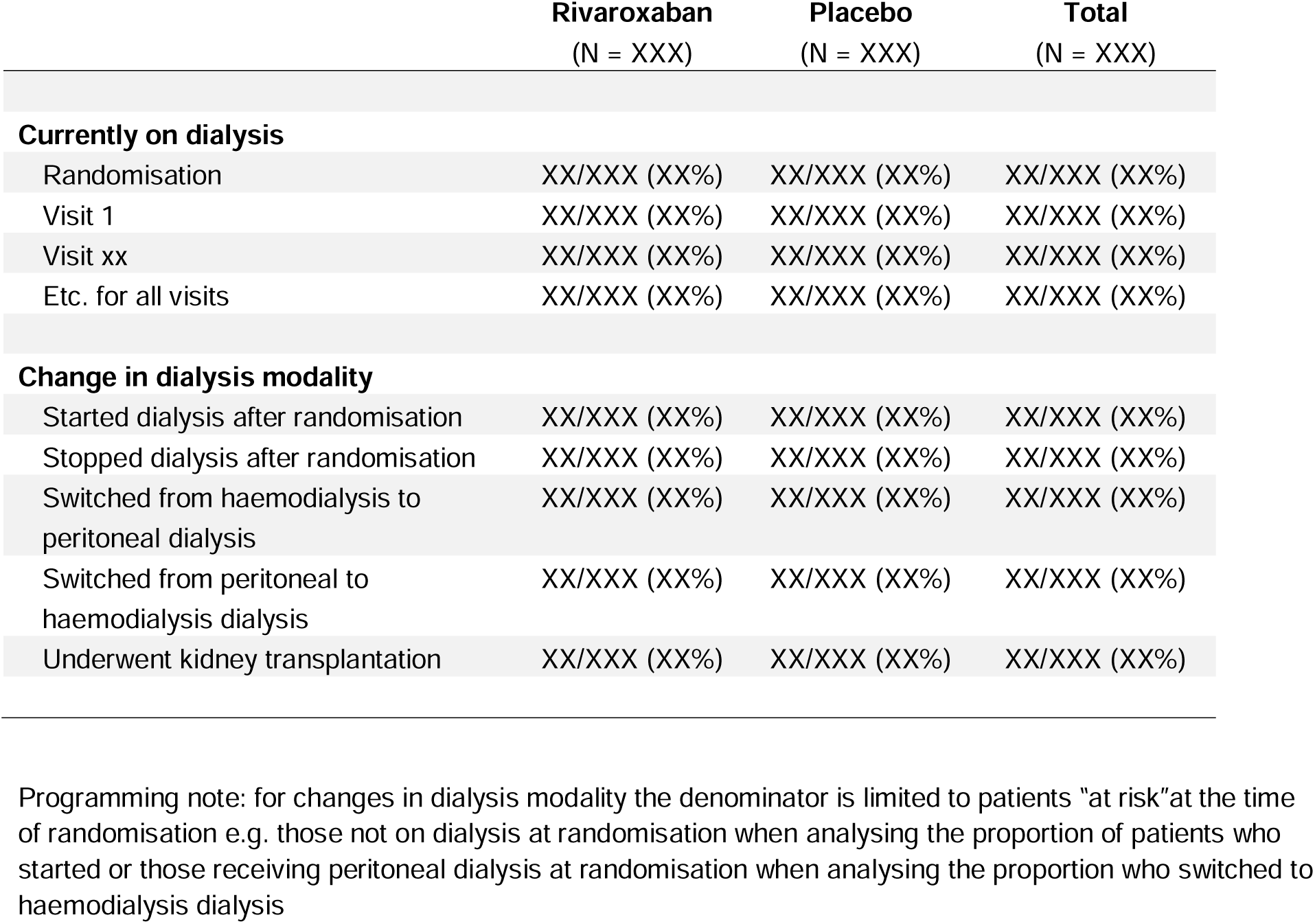
Dialysis detail.

**Table 8.** Cardiovascular and safety outcomes.

| Outcome | Rivaroxaban (N= XXX) |  | Placebo (N= XXX) |  | Difference |  |
| --- | --- | --- | --- | --- | --- | --- |
|  | N patients (%) | N events<br>(rate per 100 PY) | N patients (%) | N events<br>(rate per 100 PY) | Hazard ratio<br>(95% CI) | P-value<br>*** |
| **Primary outcome \*** | XXX (XX.X%) | XXX (XX.X%) | XXX (XX.X%) | XXX (XX.X%) | X.XX (X.XX,<br>X.XX) | X.XXX |
| **Secondary outcomes** |  |  |  |  |  |  |
| All-cause death | XXX (XX.X%) | XXX (XX.X%) | XXX (XX.X%) | XXX (XX.X%) | X.XX (X.XX,<br>X.XX) | X.XXX |
| Cardiovascular death | XXX (XX.X%) | XXX (XX.X%) | XXX (XX.X%) | XXX (XX.X%) | X.XX (X.XX,<br>X.XX) | X.XXX |
| Stroke | XXX (XX.X%) | XXX (XX.X%) | XXX (XX.X%) | XXX (XX.X%) | X.XX (X.XX,<br>X.XX) | X.XXX |
| Ischaemic or uncertain type | XXX (XX.X%) | XXX (XX.X%) | XXX (XX.X%) | XXX (XX.X%) | X.XX (X.XX,<br>X.XX) |  |
| Haemorrhagic | XXX (XX.X%) | XXX (XX.X%) | XXX (XX.X%) | XXX (XX.X%) | X.XX (X.XX,<br>X.XX) |  |
| Myocardial Infarction | XXX (XX.X%) | XXX (XX.X%) | XXX (XX.X%) | XXX (XX.X%) | X.XX (X.XX,<br>X.XX) | X.XXX |
| PAD events | XXX (XX.X%) | XXX (XX.X%) | XXX (XX.X%) | XXX (XX.X%) | X.XX (X.XX,<br>X.XX) | X.XXX |
| CV death + MI + stroke | XXX (XX.X%) | XXX (XX.X%) | XXX (XX.X%) | XXX (XX.X%) | X.XX (X.XX,<br>X.XX) |  |
| All-cause death + MI + stroke | XXX (XX.X%) | XXX (XX.X%) | XXX (XX.X%) | XXX (XX.X%) | X.XX (X.XX,<br>X.XX) |  |
| All-cause death + MI + stroke +<br>PAD | XXX (XX.X%) | XXX (XX.X%) | XXX (XX.X%) | XXX (XX.X%) | X.XX (X.XX,<br>X.XX) |  |
| Venous thromboembolism | XXX (XX.X%) | XXX (XX.X%) | XXX (XX.X%) | XXX (XX.X%) | X.XX (X.XX,<br>X.XX) |  |
|  | N patients (%) | N events<br>(rate per 100 PY) | N patients (%) | N events<br>(rate per 100 PY) | Hazard ratio<br>(95% CI) | P-value<br>*** |
| Thrombosis of dialysis access | XXX (XX.X%) | XXX (XX.X%) | XXX (XX.X%) | XXX (XX.X%) | X.XX (X.XX,<br>X.XX) |  |
| **Safety outcomes and net clinical benefit** |  |  |  |  |  |  |
| Major bleeding | XXX (XX.X%) | XXX (XX.X%) | XXX (XX.X%) | XXX (XX.X%) | X.XX (X.XX,<br>X.XX) | X.XXX |
| Gastrointestinal bleeding | XXX (XX.X%) | XXX (XX.X%) | XXX (XX.X%) | XXX (XX.X%) | X.XX (X.XX,<br>X.XX) | X.XXX |
| Net clinical benefit ** | XXX (XX.X%) | XXX (XX.X%) | XXX (XX.X%) | XXX (XX.X%) | X.XX (X.XX,<br>X.XX) | X.XXX |

Note:
\* The primary outcome is a composite of cardiovascular death, non-fatal myocardial infarction, stroke or peripheral artery disease event.
\*\* Net clinical benefit is a composite of cardiovascular death, non-fatal myocardial infarction, stroke, PAD events, fatal bleeding, or symptomatic bleeding into a critical organ
\*\*\* For 5 key secondary outcomes, p-value is adjusted using a Holm-Sidak procedure
Programming note: repeat this table for (1) per protocol analysis set (2) by aspirine treatment at baseline (yes/no) (3) dialysis status at baseline (yes/no)

**Table 9.** Major bleeding events.

| **Outcome** | **Rivaroxaban**<br>**(N= XXX)**<br>**Ev Pt/N (%)** | **Placebo**<br>**(N= XXX)**<br>**Ev Pt/N (%)** | **Fisher's**<br>**exact test**<br>**P-value \*** |
| --- | --- | --- | --- |
| **Major bleeding** | **XXX XXX/XXX (XX.X%)** | **XXX XXX/XXX (XX.X%)** | **0.xxx** |
| Fatal bleeding | XXX XXX/XXX (XX.X%) | XXX XXX/XXX (XX.X%) |  |
| Symptomatic bleeding into a critical area or bleeding into the surgical site requiring reoperation | XXX XXX/XXX (XX.X%) | XXX XXX/XXX (XX.X%) |  |
| Bleeding requiring hospitalisation | XXX XXX/XXX (XX.X%) | XXX XXX/XXX (XX.X%) |  |
| **Any gastrointestinal bleeding** | **XXX XXX/XXX (XX.X%)** | **XXX XXX/XXX (XX.X%)** | **0.xxx** |
| Overt bleeding of gastroduodenal origin confirmed by endoscopy or radiography | XXX XXX/XXX (XX.X%) | XXX XXX/XXX (XX.X%) |  |
| Overt upper gastrointestinal bleeding of unknown origin | XXX XXX/XXX (XX.X%) | XXX XXX/XXX (XX.X%) |  |
| Bleeding of presumed occult upper gastrointestinal tract origin with documented decrease in ha | XXX XXX/XXX (XX.X%) | XXX XXX/XXX (XX.X%) |  |
| **Any symptomatic bleeding** | **XXX XXX/XXX (XX.X%)** | **XXX XXX/XXX (XX.X%)** | **0.xxx** |
| Intraarticular | XXX XXX/XXX (XX.X%) | XXX XXX/XXX (XX.X%) |  |
| Intramuscular with compartment syndrome | XXX XXX/XXX (XX.X%) | XXX XXX/XXX (XX.X%) |  |
| Intraspinal | XXX XXX/XXX (XX.X%) | XXX XXX/XXX (XX.X%) |  |
| Intracranial | XXX XXX/XXX (XX.X%) | XXX XXX/XXX (XX.X%) |  |
| Intraocular | XXX XXX/XXX (XX.X%) | XXX XXX/XXX (XX.X%) |  |
| Respiratory | XXX XXX/XXX (XX.X%) | XXX XXX/XXX (XX.X%) |  |
| Pericardial | XXX XXX/XXX (XX.X%) | XXX XXX/XXX (XX.X%) |  |
| Liver | XXX XXX/XXX (XX.X%) | XXX XXX/XXX (XX.X%) |  |
| Pancreas | XXX XXX/XXX (XX.X%) | XXX XXX/XXX (XX.X%) |  |
| Retroperitoneal | XXX XXX/XXX (XX.X%) | XXX XXX/XXX (XX.X%) |  |
| Adrenal gland | XXX XXX/XXX (XX.X%) | XXX XXX/XXX (XX.X%) |  |
| Kidney | XXX XXX/XXX (XX.X%) | XXX XXX/XXX (XX.X%) |  |
| Other site | XXX XXX/XXX (XX.X%) | XXX XXX/XXX (XX.X%) |  |
| **Fatal bleeding + intracranial bleeding** | **XXX XXX/XXX (XX.X%)** | **XXX XXX/XXX (XX.X%)** | **X.XXXX** |

Notes:
1. Any bleeding includes fatal and non-fatal bleeding.
Counts displayed are number of events, number of patients / total randomised patients and percentages.
Site of bleeding includes major and non-major bleeding.
\*. P-value from the Fisher's exact test.
Programming note: repeat this table for (1) per protocol analysis set (2) by aspirine treatment at baseline (yes/no) (3) dialysis status at baseline (yes/no)

**Table 10.** EQ-5D Model results.

| **Outcome** | **Rivaroxaban**<br>**(N= XXX)**<br>**Mean (SE)** | **Placebo**<br>**(N= XXX)**<br>**Mean (SE)** | **Mean difference**<br>**(95% CI)** | **P-value** |
| --- | --- | --- | --- | --- |
| Randomisation | XX.X (X.XX) | XX.X (X.XX) | X.XX (X.XX,<br>X.XX) |  |
| Year 1 | XX.X (X.XX) | XX.X (X.XX) | X.XX (X.XX,<br>X.XX) |  |
| Year 2 | XX.X (X.XX) | XX.X (X.XX) | X.XX (X.XX,<br>X.XX) |  |
| Etc... | XX.X (X.XX) | XX.X (X.XX) | X.XX (X.XX,<br>X.XX) |  |
| **Overall** | XX.X (X.XX) | XX.X (X.XX) | X.XX (X.XX,<br>X.XX) | X.XXX |

Notes: Estimates obtained from repeated-measure linear model

**Table 11.** Summary of Adverse events.

| **Outcome** | **Rivaroxaban**<br>**(N= XXX)**<br>**Ev Pt/N (%)** | **Placebo**<br>**(N= XXX)**<br>**Ev Pt/N (%)** | **Fisher's**<br>**exact test**<br>**P-value \*** |
| --- | --- | --- | --- |
| **Any SAE** | **XXX XXX/XXX (XX.X%)** | **XXX XXX/XXX (XX.X%)** | **0.xxx** |
| Life-threatening | XXX XXX/XXX (XX.X%) | XXX XXX/XXX (XX.X%) |  |
| Required inpatient hospitalisation or<br>prolongation of existing hospitalisation | XXX XXX/XXX (XX.X%) | XXX XXX/XXX (XX.X%) |  |
| Persistent or significant disability/incapacity | XXX XXX/XXX (XX.X%) | XXX XXX/XXX (XX.X%) |  |
| Congenital abnormality/birth defect | XXX XXX/XXX (XX.X%) | XXX XXX/XXX (XX.X%) |  |
| Other important medical condition | XXX XXX/XXX (XX.X%) | XXX XXX/XXX (XX.X%) |  |

**Table 12.** Serious adverse events by system organ class.

| **Outcome** | **Rivaroxaban**<br>**(N= XXX)**<br>**Ev Pt/N (%)** | **Placebo**<br>**(N= XXX)**<br>**Ev Pt/N (%)** |
| --- | --- | --- |
| Blood and lymphatic system disorders |  |  |
| Cardiac disorders |  |  |
| Ear and labyrinth disorders |  |  |
| Eye disorders |  |  |
| Gastrointestinal disorders |  |  |
| General disorders and administration site<br>conditio |  |  |
| Hepatobiliary disorders |  |  |
| Immune system disorders |  |  |
| Infections and infestations |  |  |
| Injury, poisoning and procedural<br>complications |  |  |
| Investigations |  |  |
| Metabolism and nutrition disorders |  |  |
| Musculoskeletal and connective tissue<br>disorders |  |  |
| Neoplasms benign, malignant and<br>unspecified (incl |  |  |
| Nervous system disorders |  |  |
| Product issues |  |  |
| Psychiatric disorders |  |  |
| Renal and urinary disorders |  |  |
| Reproductive system and breast disorders |  |  |
| Respiratory, thoracic and mediastinal<br>disorders |  |  |
| Skin and subcutaneous tissue disorders |  |  |
| Social circumstances |  |  |
| Surgical and medical procedures |  |  |
| Vascular disorders |  |  |

**Table 13.** Causes of deaths.

| **Cause** | **Rivaroxaban**<br>**(N= XXX)**<br>**N (%)** | **Placebo**<br>**(N= XXX)**<br>**N (%)** |
| --- | --- | --- |
| **Cardiovascular death** | **XXX (XX.X%)** | **XXX (XX.X%)** |
| Acute myocardial infarction | XXX (XX.X%) | XXX (XX.X%) |
| Sudden cardiac death | XXX (XX.X%) | XXX (XX.X%) |
| Heart failure | XXX (XX.X%) | XXX (XX.X%) |
| Stroke | XXX (XX.X%) | XXX (XX.X%) |
| Cardiovascular procedure | XXX (XX.X%) | XXX (XX.X%) |
| Other cardiovascular cause | XXX (XX.X%) | XXX (XX.X%) |
| **Non-cardiovascular death** | **XXX (XX.X%)** | **XXX (XX.X%)** |
| Infection | XXX (XX.X%) | XXX (XX.X%) |
| Renal | XXX (XX.X%) | XXX (XX.X%) |
| Gastrointestinal/Hepatobiliary/Pancreatic | XXX (XX.X%) | XXX (XX.X%) |
| Malignancy | XXX (XX.X%) | XXX (XX.X%) |
| Pulmonary | XXX (XX.X%) | XXX (XX.X%) |
| Fatal bleeding | XXX (XX.X%) | XXX (XX.X%) |
| Other | XXX (XX.X%) | XXX (XX.X%) |
| **Undetermined** | **XXX (XX.X%)** | **XXX (XX.X%)** |

### 4.5 Analysis of the primary outcome

The primary outcome is a composite of death from cardiovascular causes, myocardial infarction, stroke or peripheral artery disease events. The main analysis will consist of a survival analysis of time to first event conducted with a Cox proportional model. Participants will be censored at the time when they die from a cause that is not cardiovascular. Censoring will otherwise occur when the participant was last known to be alive and free of event. Patients undergoing kidney transplantation had their study medication stopped but still remained in the trial; therefore, the survival analysis will not be censored at the time of kidney transplantation unless this coincides with the time when the patient was lost to follow-up (i.e. last known to be alive and event-free). As a sensitivity analysis, death from non-cardiovascular causes will be treated as competing risks (see Section 4.5.2.2 for details).

#### 4.5.1 Main analysis of the primary outcome

Survival will be described using Kaplan-Meier curves of time-to-first event. The effect of the intervention will be estimated as a hazard ratio and confidence interval from a Cox model. Statistical significance will be assessed using the p-value from the Cox model associated with the hazard ratio. To model potential within-site correlations due to stratification, we will use a shared-parameter frailty Cox model with a random site effect [4]. Fixed covariates will include the randomised treatment and the other 3 stratification variables (use of aspirin, CKD stage and diabetes). Raw event rates (per 100 patient-years) and 95% confidence intervals will be calculated using a Poisson model.

#### 4.5.2 Sensitivity analyses

##### 4.5.2.1 Adjusted analyses

The Cox model described in Section 4.5.1 will be rerun after adding the following baseline covariates to the model: age (as a continuous variable), sex,smoking status and region ((1)India and Nepal, (2) SE Asia, (3) Saudi Arabia and Tunisia, (4) Australia, Europe and Canada). Additional adjustments may be considered in case of substantial imbalances in baseline characteristics (absolute standardised difference > 0.1) not already included as covariates.

##### 4.5.2.2 Competing risk analysis

The model described in Section 4.5.1 will be rerun by treating death from non-cardiovascular causes as a competing risk. Survival will be summarized using cumulative incidence functions and the effect of the intervention will be derived from a Fine-Gray model as the sub-distribution hazard ratio and 95% confidence interval [5].

##### 4.5.2.3 Subgroup analyses

The following subgroup analyses are planned for the primary outcome, regardless of the statistical significance for the primary analysis:

- Age: <65, 65 or more
- Sex: male vs. female
- Country (grouped as (1) India and Nepal, (2) SE Asia, (3) Saudi Arabia and Tunisia, (4) Australia, Europe and Canada)
- Aspirin use: yes vs no
- Diabetes mellitus: yes vs no
- End-Stage Kidney Disease (receiving dialysis): yes vs no
- Smoking status: current or previous smokers vs never smoked

Subgroup analyses will be performed by adding the relevant subgroup variable as well as its interaction with the randomised intervention to the main analysis model described in Section 4.5.1. The p-value associated with the interaction term will be used to assess potential heterogeneity of intervention effect by subgroups.

### 4.6 Analysis of other outcomes

#### 4.6.1 Analysis of secondary outcomes

All secondary outcomes (see Section 3.3.2) will be analysed using the same approach as the primary outcome described in Section 4.5.1 i.e. described using Kaplan-Meier plots and using a Cox model of time to first event with adjustment for stratification variables to estimate the effect of the intervention. Except where the outcome itself is or includes all-cause mortality, death will be treated as a censoring mechanism. For all-cause death, we will apply the same adjustments and subgroup analyses as for the primary outcome (see Sections 4.5.2.1 and 4.5.2.3). No adjustment or subgroup analysis will be performed on other secondary outcomes.

#### 4.6.2 Analysis of tertiary outcomes

Thrombosis of dialysis vascular access among participants with an AV fistula/graft will be using the same approach as the primary outcome described in Section 3.3.1 i.e. described using KM plots and using a Cox model of time to first event with adjustment for stratification variables to estimate the effect of the intervention. Death will be treated as a censoring mechanism. Only participants with an AV fistula/graft at baseline will be included in the denominator.

Each of the five EQ-5D-5L dimensions will be described by visit using bar charts. The visual analogous scale ratings will be described using a box plot and summarised as the mean, standard deviation, median and quartiles. The visual analogous scale (score of 0 to 100) will be analysed using linear model (i.e. assuming a normal distribution and an identity link function) which will include all post-randomisation measurements. The following variables will be included as fixed effects: the randomised arm, the baseline visual analogous scale value, the visit and the interaction between visit and treatment. Visit will be considered as a categorical variable thus not assuming any specific trend/shape over time. Correlations between repeated measures (within subject) will be modelled using a repeated effect assuming an exchangeable correlation matrix. The effect of the intervention will be estimated as the overall mean difference and 95% CI.

Further analyses of the EQ-5D-5L, including health utility scores, will form part of a separate health economic analysis.

#### 4.6.3 Analysis of safety outcomes

Major bleeding events will be summarised as the number of events as well as the number and proportion of patients experiencing at least one event. This will be done overall, by outcome and by site of bleeding. Fisher’s exact test will be used to test differences in proportions.

Time to first major bleeding event will be described by randomised arm using Kaplan-Meier plots and analysed using a Cox model adjusted for stratification variables using the same approach as the analysis of the primary efficacy outcome (see Section 4.5.1). Censoring will occur when the patient was last known to be alive and free of major bleeding event. The same analysis will be conducted for gastrointestinal bleeding events and for the net benefit outcome (as defined in Section 3.3.5).

These analysed will be repeated in the per-protocol analysis population and by baseline use of aspirin and baseline dialysis status.

#### 4.6.4 Win ratio analysis

All-cause death and components of the primary outcome will be analysed jointly using a win ratio approach which ranks all outcomes in order of importance [6]. The following hierarchy will be used:

- Death due to any cause
- Non-fatal stroke
- Non-fatal myocardial infarction
- Non-fatal peripheral artery disease event

Every participant randomised to the rivaroxaban arm will be paired with every participant randomised to the placebo arm. For illustrative purposes, if there were 130 patients randomised to one arm and 127 to the other, this approach would compare a total of 16,510 pairs (130 × 127). Within each pair, outcomes will be compared in a hierarchical fashion until a winner has been determined. If no winner can be declared after comparing all outcomes, the pair will be tied. This will start by comparing death due to any cause to determine a “winner”. The winner will be the participant with the longest survival time. If neither participant died and a winner cannot be declared on mortality alone, it will then proceed to the next step and compare time to stroke. As with death, the winner will be the participant with the longest time to first event. If neither patient experienced the event of interest or both experienced the event on the same day, the pair will be declared a tie and the analysis will proceed to the next outcome in the hierarchy. Only events that occur during the time that both participants are in the study will be used (e.g. if one participant withdraws after 1 year, only the events occurring in the first year will be used to compare the pair). The approach will proceed in a stepwise fashion, moving to the next outcome in the hierarchy, until a decision has been reached for every pair. The full decision process is described in the table below. The win ratio will then be calculated as the proportion of “rivaroxaban winners” (where the participant in the rivaroxaban arm had a better outcome) divided by the proportion of “placebo winners” (where the participant in the placebo arm had a better outcome)

To reduce the potential impact of confounding and potentially improve statistical efficiency, the win ratio approach will be stratified by country following the method proposed by Dong et al. [4]. This means that patients from the rivaroxaban arm from one country will only be paired (and compared) to patients from the placebo from the same country. To complete the interpretation, we will also compute the stratified win odds and net benefit [8,9]. Confidence intervals will be computed using the delta method as described by Dong et al. [4]. Calculations will be performed using the R package WINS developed by Cui and Huang [10].

| Step | Outcome | Decision rule |
| --- | --- | --- |
| 1 | Time to all-cause death | The winner is the patient with the longest survival time. If neither died or if both died on the same day, go to step 2. |
| 2 | Time to first non-fatal stroke | The winner is the patient with the longest time to event. If neither patient experienced the event or both patients had it on the same day, go to step 3. |
| 3 | Time to first non-fatal MI | The winner is the patient with the longest time to event. If neither patient experienced the event or both patients had it on the same day, go to step 4. |
| 4 | Time to first non-fatal PAD event | The winner is the patient with the longest time to event. If neither patient experienced the event or both patients had it on the same day, the comparison stops and the pair is considered a tie. |

An “incremental analysis” will also be performed by calculating the win ratio and 95% CI after adding each outcome i.e. for the first outcome only, for the first two together, the first three, and so on (cf Figure 4 for details).

**Figure 1.**
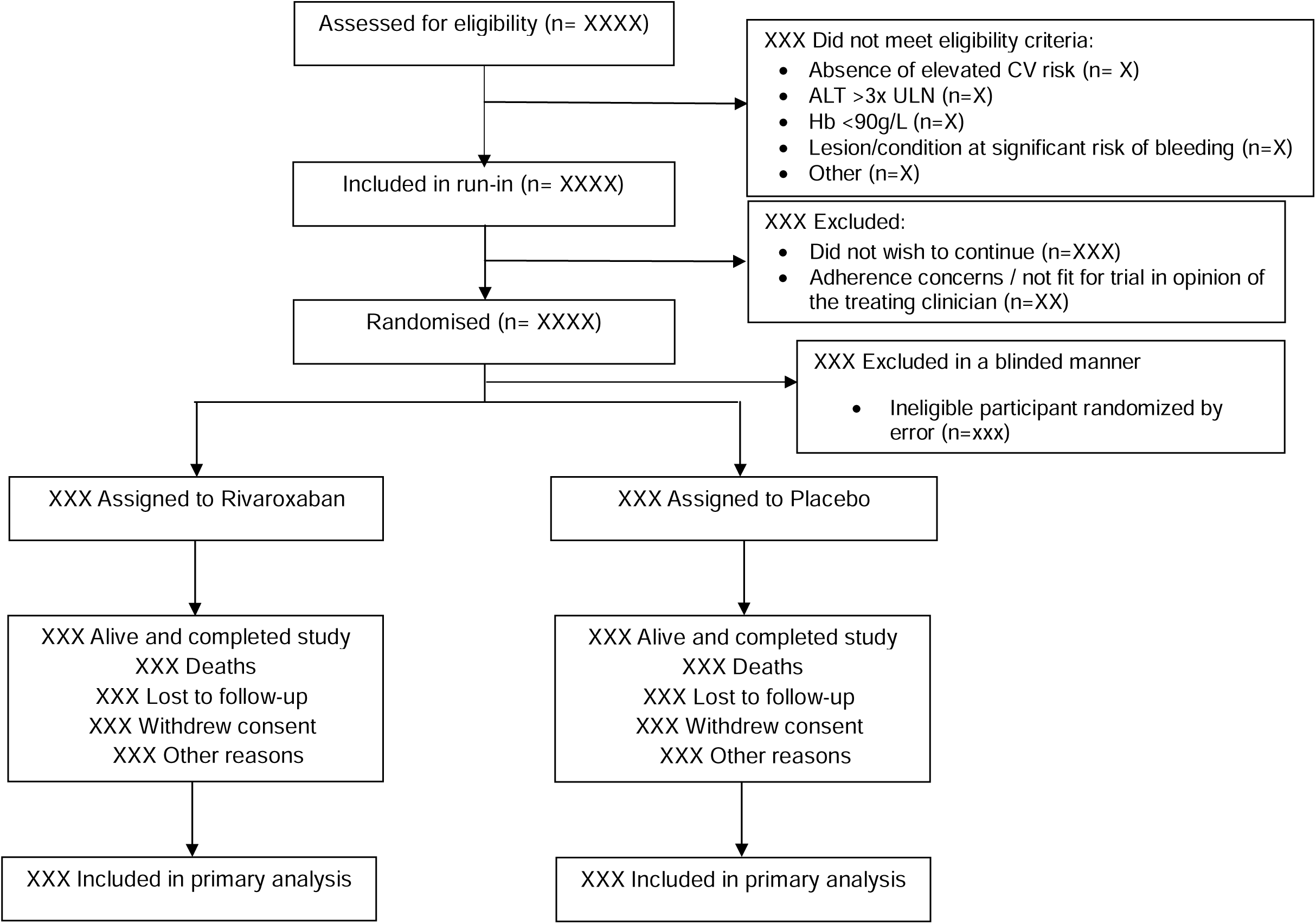
Consort flow chart

**Figure 2.**
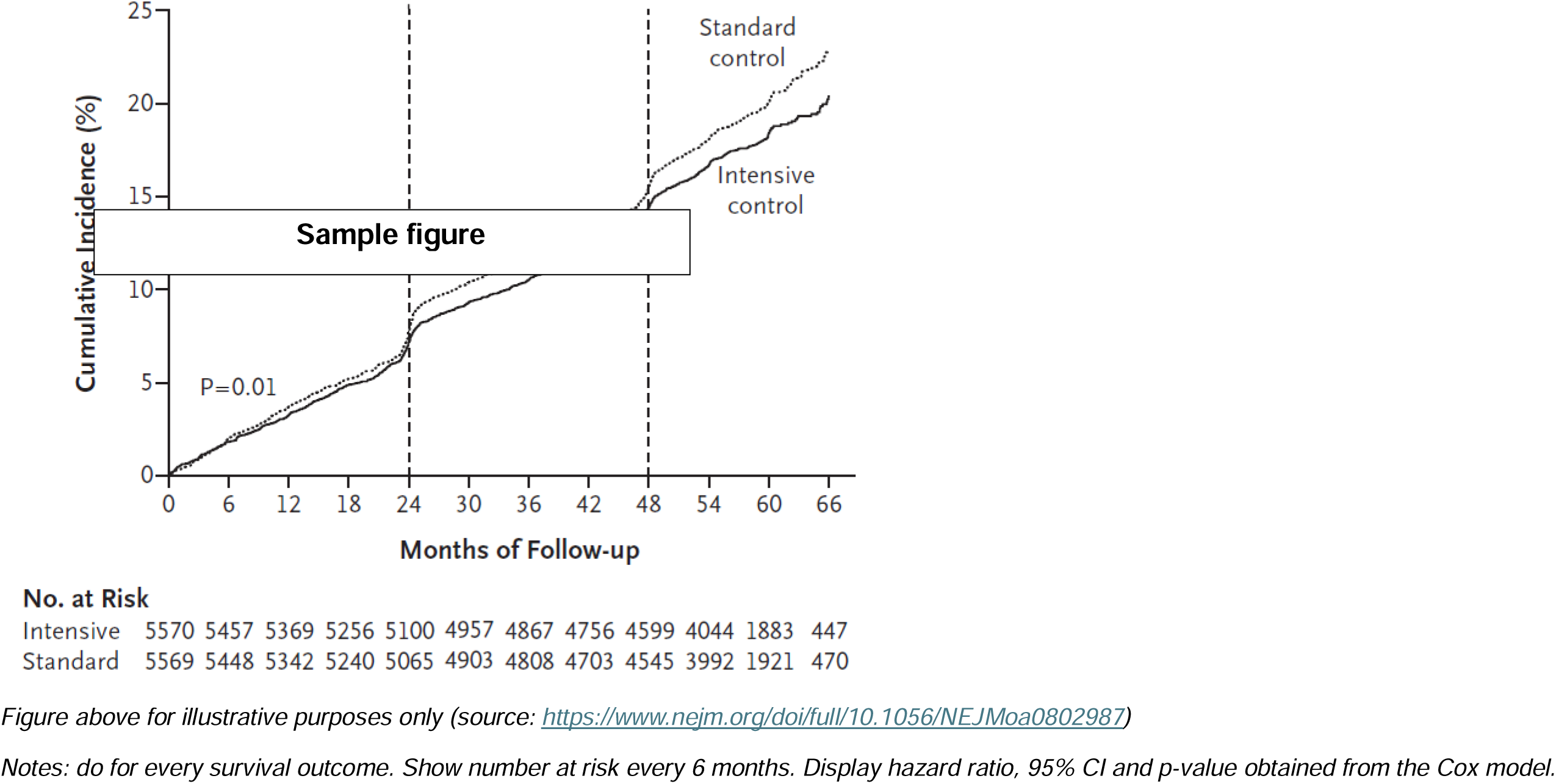
Kaplan-Meier plot of time to first primary outcome

**Figure 3.**
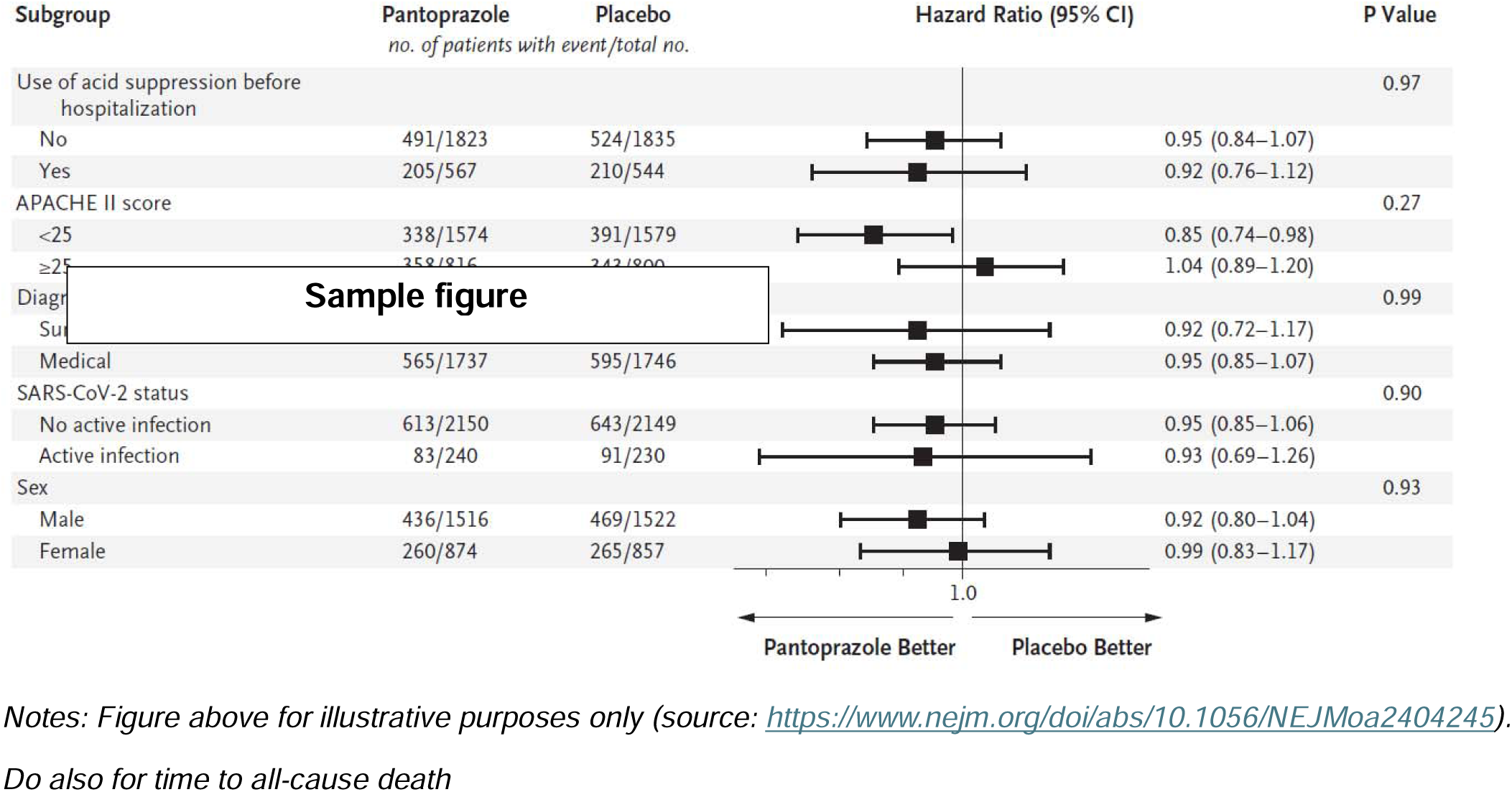
Forest plot of subgroup analyses of the primary outcome

**Figure 4.**
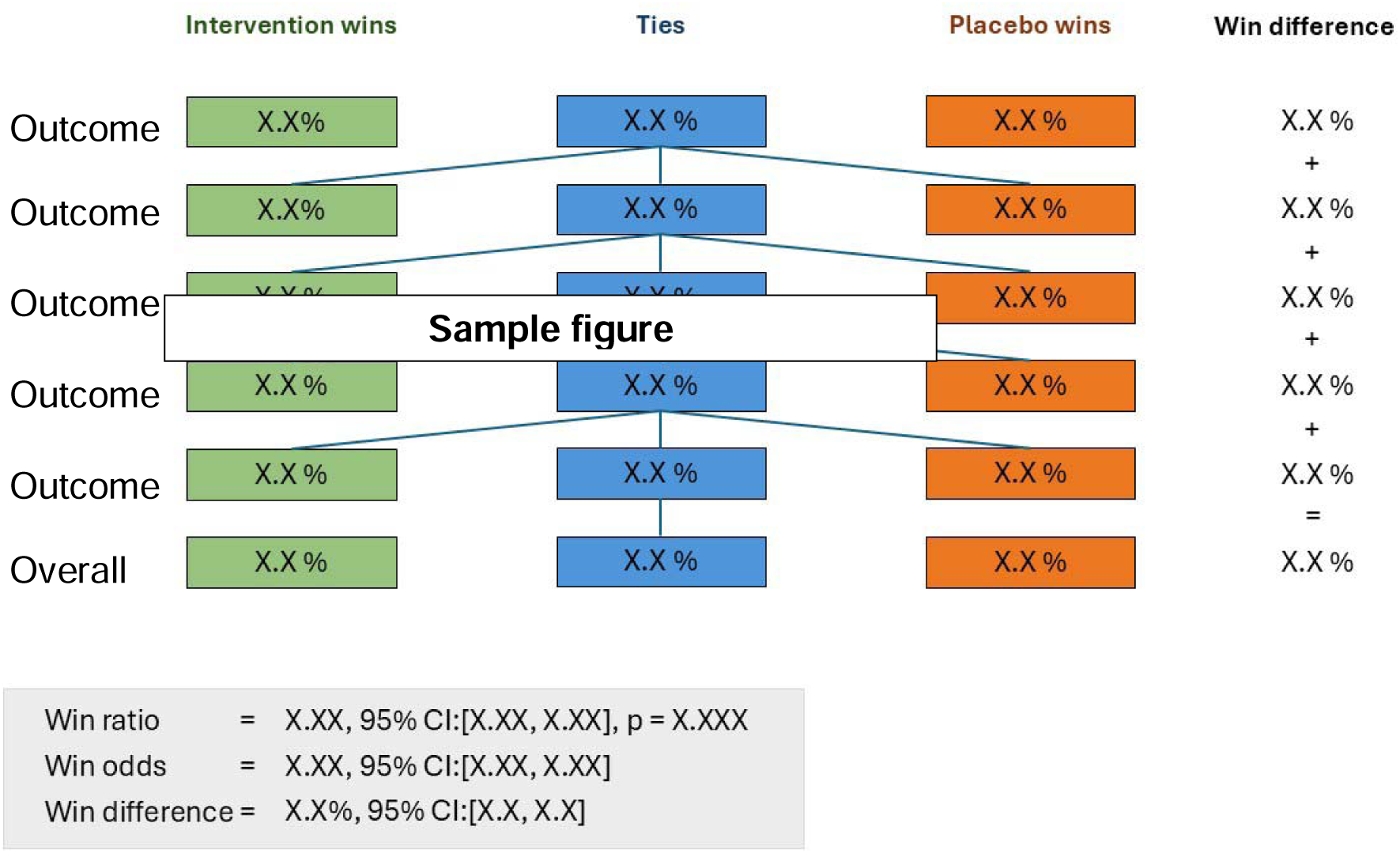
Win ratio decision tree

**Figure 5.**
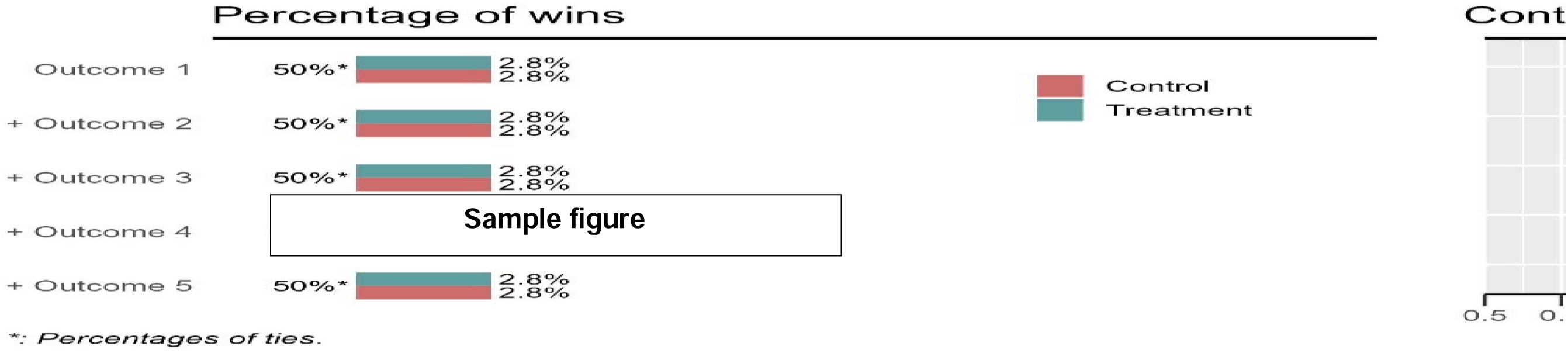
Win ratio cumulative analysis

**Figure 6.** Boxplot of EQ-5D VAS by visit. Programming note: show boxplot for each arm side by side with a panel per visit

**Figure 7.**
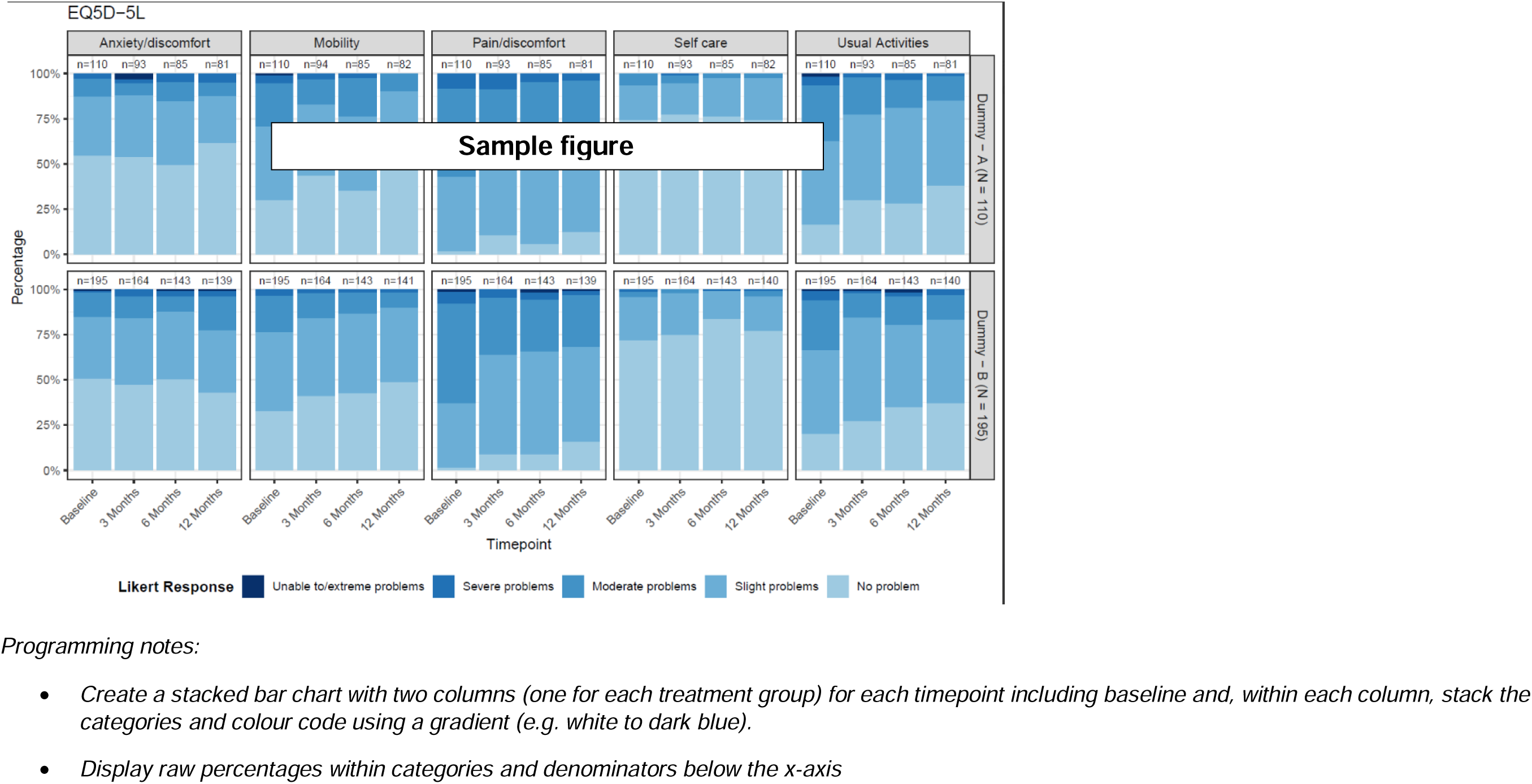
Stacked bar chart of EQ5D-5L by visit

## Data Availability

Not relevant.

## 2 Administrative information

### 2.1 Study identifiers

- Protocol version: v4.0 24 Feb 2022
- ClinicalTrials.gov register Identifier: NCT03969953

### 2.2 Contributors to the statistical analysis plan

#### 2.2.1 Roles and responsibilities

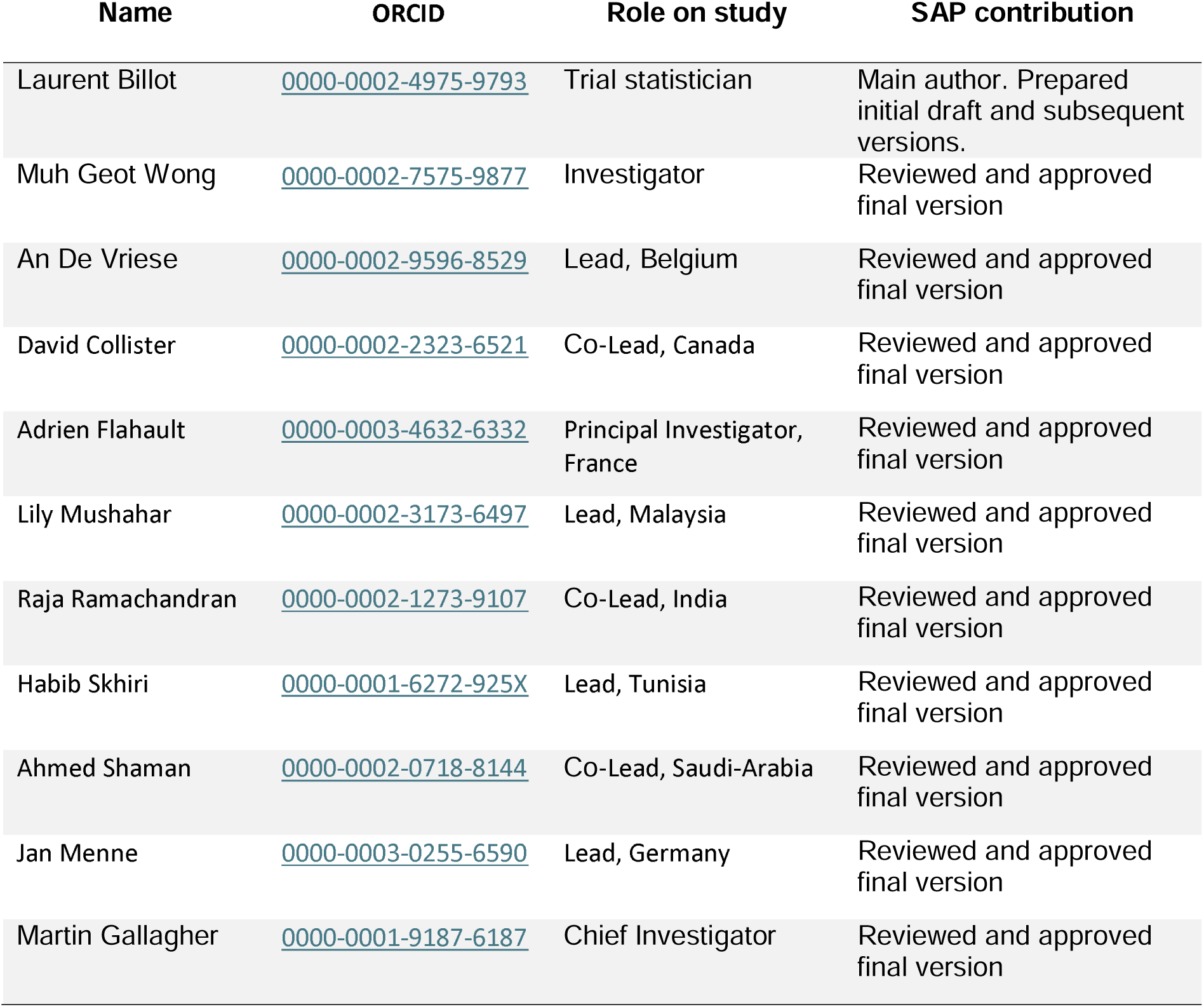

#### 2.2.2 Approvals

The undersigned have reviewed this plan and approve it as final. They find it to be consistent with the requirements of the protocol as it applies to their respective areas. They also find it to be compliant with International Conference on Harmonisation (ICH-E9) principles and in particular, confirm that this analysis plan was developed in a blinded manner (i.e. without knowledge of the effect of the intervention being assessed).

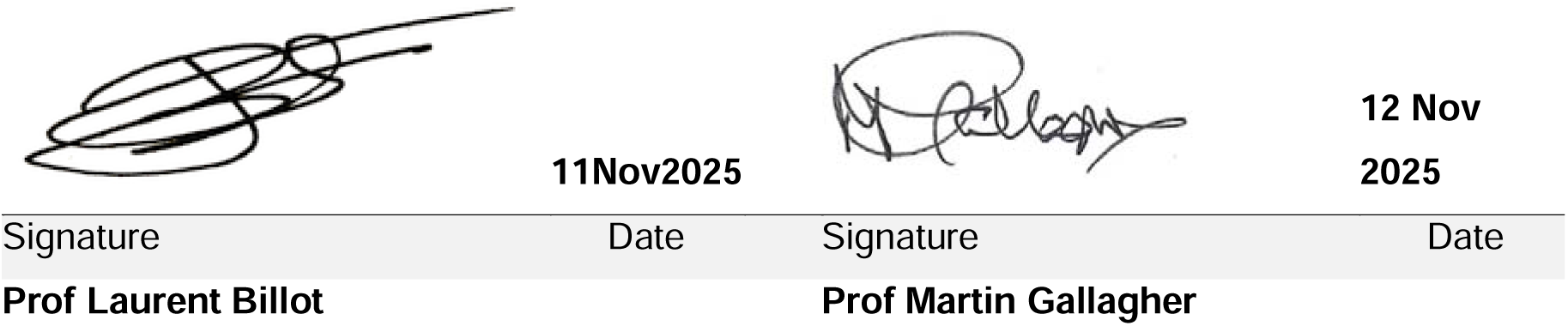

## Notes

### Competing Interest Statement

The authors have declared no competing interest.

### Clinical Trial

NCT03969953

### Funding Statement

The TRACK trial is funded by grants from the National Health and Medical Research Council of Australia (2018/GNT1162375), the French Ministry of Health Programme Hospitalier de Recherche Clinique (Projet PHRC-19-0112) and Deutsche Forschungsgemeinschaft, Germany (Project number 506165771). Rivaroxaban 2.5 mg tablets and placebo tablets were provided by Bayer AG, Germany free of cost through the
Investigator Initiated Research Support Scheme.

### Author Declarations

Full ethic approval as been obtained from the following committees: - Australia: South Eastern Sydney Local Health District HREC - Belgium: Commissie voor Ethiek AZ Sint-Jan Brugge-Oostende AV - Canada: Hamilton Integrated Research Ethics Board & Health Research Ethics Board - Biomedical Panel - France: Comite de Protection des Personnes Ouest VI - Germany: Ethikkommission der Universitat zu Lubeck - India:The George Institute Ethics Review Committee - Malaysia: Central Ethics Medical Research and Ethics Committee & Universiti Kebangsaan Malaysia Research Ethics Committee & Medical Research Ethics Committee, University Malaya Medical Centre - Nepal: Tribhuvan Institute of Medicine Research Committee - Singapore: National Health Group Domain Specific Review Board (NHG DSRB) - Saudia Arabia: KAIMRC Institutional Review Board & King Saud University College of Medicine Institutional Review Board - Taiwan: Institutional Review Board of Fu Jen Catholic University Hospital - Tunisia: Comite de Protection des Personnes du Centre CPP

